# A standardised saffron extract improves subjective and objective sleep quality in healthy older adults with sleep complaints: results from the Gut-Sleep-Brain Axis randomised, double-blind, placebo-controlled pilot study

**DOI:** 10.1101/2025.02.20.25322405

**Authors:** Leonie Lang, Annabel Ditton, Andi Stanescu, Vernisse Jainani, Simon McArthur, Line Pourtau, David Gaudout, Matthew G. Pontifex, Jordan Tsigarides, Taylor Steward, Saber Sami, Michael Muller, Michael Hornberger, David Vauzour, Alpar S. Lazar

**Author notes:** These authors share senior authorship (David Vauzour, Alpar S. Lazar).

## Abstract

Sleep disorders may increase neurodegenerative diseases risks and alter gut microbiota composition. Saffron (*Crocus sativus*) supplementation has been linked to sleep improvements and gut microbiome changes, but its effect on sleep quality through gut microbiota-brain axis modulation remains unexplored. A randomised, placebo-controlled pilot study of saffron (30mg/day for 4-weeks) was conducted with 52 older adults (55-85 years) experiencing sleep complaints. Subjective and objective sleep quality were measured using validated questionnaires and electroencephalography-based sleep tracker respectively. A subgroup (n=26) underwent microbiome analysis. Saffron supplementation improved subjective sleep quality (p=0.02) and sleep efficiency (p=0.04). Objective measures showed reduced latency to persistent sleep (p=0.003) and sleep onset latency (p=0.03). Linear discriminant analysis effect size analysis of the microbiome revealed increases in *Faecalibacterium* (q=0.013), *Lachnoclostridium* (q=0.045), *Prevotella* (q=0.022), *UBA1819* (q=0.020) and *Oscillibacter* (q=0.045), while decreasing *Dialister* (q=0.028). Univariate analysis further indicated increases in *Lachnospiraceae-UGC-001* (p=0.020) and *Roseburia* (p=0.03), with a decrease in *Turicibacter* (p=0.045) in the saffron group. *Oscillibacter* and *UBA1819* correlated with subjective measures of sleep efficiency (r=0.63, p=0.0007) and sleep latency (r=-0.39, p=0.04. Changes in *Dialister, Turicibacter* and *UBA1819* were correlated with several measures of objective sleep quality including wake duration, latency to persistent sleep and wake-after-sleep-onset. Four-weeks saffron supplementation improved both subjective and objective sleep quality in older adults with sleep complaints, while increasing gut bacteria that produce short-chain fatty acids. These results pave the way for further randomised controlled trials exploring the links between sleep quality and gut health and may help devising new preventative strategies for age-related brain disorders.

## 1. Introduction

The global population is ageing, with the number of people over 60 expected to double by 2050. ^1^ As ageing is a major risk factor for neurodegenerative conditions such as dementia ^2^, the burden of such diseases is similarly expected to rise in the coming years. Intriguingly, ageing is associated with an increased prevalence of sleep deficits ^3, 4^ which may represent one of the contributing factors to neurodegeneration. ^5, 6^ Indeed, sleep plays a key role in multiple physiological functions including regulation of neural cellular energy homeostasis, neural plasticity and clearance of brain waste products. ^7–9^ However, with age, physiological sleep patterns deteriorate ^4, 10–12^ with reduction in slow wave sleep, rapid-eye-movement (REM), sleep efficiency and sleep continuity. The causality of such age-related sleep problems is complex, and to date there are no effective, widely available interventions to improve sleep deficits in the elderly. ^13^ Given the significance of sleep for brain health, targeting sleep quality improvement in older adults has a significant potential to improve the quality of life and to potentially mitigate factors associated with neurodegenerative disorders in later life. ^14^

The gastrointestinal tract is now well recognised as a powerful modifier of brain function ^15, 16^ through its bidirectional communication with the brain, referred to as the microbiota gut-brain-axis. ^17^ Evidence supports a regulatory role for the gut microbiome in sleep homeostasis (e.g. converting substrates into the neurotransmitters required for sleep regulation or their precursors ^18^ or acting as signalling molecules ^19^) and that alterations in sleep duration and quality influence the composition and diversity of the gut microbiome. ^20^ Disturbances in gut eubiosis is observed in people with insomnia and circadian misalignment. ^21^ In addition, partial sleep deprivation also alters the composition of the gut microbiome, and global sleep deprivation has widespread negative effects on cognitive performance including non-spatial memory, working memory, cognitive and emotional functioning in both animal models ^22, 23^ and human studies. ^24–26^ However, whether positive sleep changes may be modulated by reshaping the gut microbiome currently remains unexplored.

Diet modulates both sleep hygiene and the gut microbiota with bioactive compounds naturally present in foods playing a fundamental role in sleep quality and in the maintenance of gut homeostasis through the production of gut-derived metabolites such as short-chain fatty acids or signalling molecules (e.g., GABA, serotonin, indoles). ^27^ Saffron (from *Crocus sativus L.*) is a spice rich in carotenoid-related compounds namely crocetin, crocin, picrocrocin, and safranal, and has been reported to positively affect sleep in both animal models and human trials. ^28–30^ The proposed direct and indirect mechanisms of saffron on sleep seem to act on the production of melatonin via the serotonin pathway or affect the levels of neurotransmitters involved in sleep regulation via the serotonergic, glutamatergic or GABAergic systems. ^31^ In preclinical studies, in addition to improving non-rapid eye movement (NREM), saffron was recently associated with beneficial shifts in the gut microbiota. ^32^ In humans, a saffron extract (15.5mg per day) for 6 weeks improved sleep quality, reduced sleep latency and improved sleep duration on the Pittsburgh Sleep Quality Index (PSQI). ^29^ Furthermore, crocetin, an active constituent of saffron, at 7.5mg/day for 2 weeks increased electroencephalography delta activity in healthy adult participants with mild sleep complaints. ^33^ However, the effect of saffron on sleep quality through the modulation of the gut microbiota-brain axis has not yet been investigated.

The main objective of this study was to investigate the impact of daily saffron extract intake (30 mg/day for 4 weeks) on both subjective and objective sleep quality in healthy older adults with sleep complaints. In addition, we also sought to assess the impact of saffron supplementation on gut microbiota diversity and abundance, and to establish whether gut microbial changes may be related to improvements in both self-reported and objectively measured sleep outcomes.

## 2. Methods

### 2.1. Ethical approval

The conduct, evaluation and documentation of this study abide with the Good Clinical Practice (GCP) guidelines and the principles of the Declaration of Helsinki. The study protocol was approved by the UEA FMH Ethics Committee (ETH2122-1829). All participants provided signed informed consent prior to participating to the study. The trial was registered at https://clinicaltrials.gov (NCT05315986).

### 2.2. Study design and participants

The “Gut-Sleep-Brain axis” trial was a four-week, double-blind, randomised, placebo-controlled pilot study conducted at a single site in Norwich, UK. Participants were recruited from an internal database of the Sleep and Brain Research Unit (SBRU) at the University of East Anglia, and via the Joint Dementia Research Forum (https://www.joindementiaresearch.nihr.ac.uk). Following recruitment and written informed consent, participants were screened using online questionnaires. If deemed eligible participants were asked to attend 2 clinical visits, at 0 weeks (baseline) and 4 weeks to undergo subjective measures and to collect/dispose sleep devices and biological samples. Microbiota speciation analysis was conducted at baseline and 4 weeks in a subset of n=26 participants. The study started in April 2022 and was completed in November 2022.

Participants were aged ≥ 55y and presented with sleep complaints as indicated by high scores on the Pittsburgh Sleep Quality Index (PSQI; > 5) or the Insomnia Severity Index (ISI; > 10) but were otherwise of a normal cognitive status (ACE-III score > 21). ^34^ Participants were excluded if they had a BMI ≥35 kg/m², were pregnant, had current neurological or psychiatric conditions (e.g., anxiety, depression, PTSD), acute infections, or chronic medical conditions that interfere with sleep (e.g., pain, tumours), a diagnosis of untreated sleep apnoea or another sleep-related disorder (excluding insomnia), were shift workers, were using antibiotics or dietary supplements that may affect sleep or sleep medications, consumed excessive caffeine (>5 cups per day of caffeinated beverages) or alcohol (>14 units/week), used nicotine or recreational drugs, or had previously reported allergies to saffron. In addition, other baseline characteristic measures were used to screen for depression and anxiety including the Patient Health Questionnaire (PHQ-9), with the exclusion of participants with high scores (>19) ^35^; the Generalised anxiety disorder (GAD), excluding participants with scores >14 ^36^, the General Health Questionnaire (GHQ-28), used to screen for psychological well-being ^37^, as well as the Positive and Negative Apathy Scale (PANAS) ^38^ to assess positive and negative affect.

### 2.3. Intervention, assignment to groups and blinding

Participants were randomly allocated to receive 30mg saffron extract (Safr’Inside^TM^, Activ’Inside, France) standardised to contain crocins [(mainly trans-4-GG, trans-3-Gg; cis-4-GG, trans-2-G) > 3%, safranal > 0.2%, picrocrocin derivatives (mainly picrocrocin, HTCC) > 1%, and kaempferol derivatives (mainly kaempferol-3-sophoroside-7-glucoside, kaempferol-3-sophoroside) >0.1%, measured by UHPLC method)] or a placebo. These compounds are natural carotenoid-type and flavonoids found in the stigmas of the saffron (*Crocus sativus L.*) flowers, providing a typical red color of specific interest for various industries ^39^, in addition to their potent pharmacological effects. ^40,41^ The 30 mg/day dose was determined based on previous randomised, double blind placebo-controlled studies reporting improvements in psychological stress response and reduced depression.^42,43^

Treatments were provided in form of gummies and participants were asked to consume one gummy every day, 30 minutes before bedtime for 4 weeks. Each gummy contained pectin, sorbitol, maltitol, stevia, orange aromas, citric acid, and sodium citrate with 30mg Safr’Inside^TM^ for the active group or without for the placebo. All the ingredients used for the manufacture of gummies were of food grade and manufactured in a FSSC 22000 certified facility.

Treatments were fully randomised by an automated randomisation algorithm, using the random size block randomisation system via an online randomisation platform (Sealed envelope, 2021). All researchers, clinical staff and participants were blinded to the treatment until all data and samples had been analysed according to a pre-defined statistical analysis plan. Adherence to the supplementation intervention was evaluated through daily compliance assessment forms filled out by participants and returned at the end of the trial, and by monitoring gummy containers returned at the end of the study.

### 2.4. Subjective and objective sleep

The primary outcome of the study was self-reported sleep quality as measured by the PSQI at baseline and post-intervention. The PSQI is a clinically validated questionnaire consisting of 19 individual items grouped into seven components, with higher scores indicating poorer sleep quality (from 0 to 21). ^44^ Six of the seven components were included in the analysis, with the exception of the (sleep) medication use component, as this was an exclusion criterion. In order to better match with the objective measurement of sleep efficiency expressed as a percentage (%), the sleep efficiency subscore of the PSQI has also been expressed in %, with higher values representing better sleep quality. Additionally, the ISI, which measures both nighttime and daytime components of insomnia (composite score from 0 to 28) ^45^ along with the Epworth Sleepiness Scale (ESS) ^46^, which assesses habitual daytime sleepiness (composite score from 0 to 24), and the single-item Karolinska Sleepiness Scale (KSS) ^47^, which measures actual sleepiness on a 9-point Likert scale, were used to characterise subjective sleep. Higher scores reflect greater levels of insomnia, habitual daytime sleepiness, and actual sleepiness, respectively.

Objective sleep was recorded for 7 consecutive nights at both baseline and post-intervention using the Dreem 3 headband (DH, Dreem 3, Beacon Biosignals, Inc). The DH is composed of foam and lightweight fabric, adjustable via an elastic band to minimise discomfort but sufficiently tight to be secure. It collects signals via 5 dry EEG sensors made of silicone with soft and flexible projections, positioned on the forehead (2 frontal, 1 ground and 2 occipital sensors), capturing at a sampling rate of 250 Hz. Supplementary details were published earlier. ^48^ Sleep diaries were used to complement objective sleep data. The sleep staging was generated for each 30-second epoch during the night, and the sleep features were generated from the raw data using an artificial neural network algorithm selecting the channels with the highest quality (referred to as the virtual channels ^49^), to provide the automatic sleep staging. The algorithm processes data in 30-second epochs to provide accurate and reliable sleep scoring. The automatically analysed sleep features included Total Sleep Time (TST, in minutes), Sleep Onset Latency (SOL, in minutes) – the duration from lights off to falling asleep – Wakefulness After Sleep Onset (WASO, in minutes), which measures the time spent awake between sleep onset and final awakening, and Latency to Persistent Sleep (LtPS, in minutes), defined as the time from eye closure to the first sustained period of non-wakefulness. Additionally, the analysis covered the time spent in NREM sleep and its sub-stages (N1, N2, and N3, in minutes), in REM sleep (in minutes), sleep efficiency (the ratio of time spent asleep to total recording time), and the number of awakenings, which indicates the number of transitions from different sleep stages to wakefulness during the night (in minutes). The normalised sleep architecture was also returned (i.e., time spent in a certain sleep stage relative to TST, in percentage). The automatic analysis of sleep (i.e., sleep scoring and extraction of sleep parameters) followed the recommendations of the American Academy of Sleep Medicine (AASM) ^50, 51^.

### 2.5. Gut microbiome

Faecal samples were collected up to 48 hours prior to treatment onset and again in the 48 hours prior to follow-up visits using the Easy Sampler collection kits (GP Medical Device, Holstebro, Denmark). Containers were stored in a cool, dry location prior to returning to the research facility at the earliest and transferred at -80°C for further analysis. Microbial DNA was isolated from approximately 50mg of faecal content using the QIAamp PowerFecal Pro DNA Kit (Qiagen, Manchester, UK) as per the manufacturer’s instructions. DNA quantity was assessed using a Nanodrop 2000 Spectrophotometer (Fisher Scientific, UK), and quality was established using agarose gel electrophoresis to detect DNA integrity, purity, fragment size and concentration. The 16S rRNA amplicon sequencing of the V3–V4 hypervariable region was performed with an Illumina NovaSeq 6000 PE250. Sequence analysis was performed by Uparse software (Uparse v7.0.1001), using all the effective tags. Sequences with ≥97% similarity were assigned to the same OTUs. A representative sequence for each OTU was screened for further annotation. For each representative sequence, the Mothur software was used to perform each sequence against the SSUrRNA database of SILVA Database 138. ^52^ OTUs abundance was normalised using a standard of sequence number corresponding to the sample using the least sequences. Alpha-diversity was assessed using standard metrics (e.g., Chao1 and Shannon H diversity index) while beta divergence was assessed using Bray-Curtis. Statistical significance was determined by Kruskal–Wallis or Permutational Multivariate Analysis of Variance (PERMANOVA) ^53^ combined with a false discovery rate (FDR) approach used to correct for multiple testing.

### 2.6. Dietary assessment

Participants were asked to fill in the Scottish collaborative group food frequency questionnaire (SCG-FFQ; version 6.6), a validated, semi-quantitative dietary assessment instrument that has been developed to estimate and rank the dietary intake of a wide range of nutrients in large-scale UK epidemiological studies. ^54, 55^ The SCG-FFQ covers 169 food items grouped into 21 categories (e.g. breads and breakfast cereals) and is described elsewhere. ^56^ It was used to describe each participant’s habitual diet over the previous two to three months, a key complementary outcome to the gut microbiome baseline measures. Participants completed the paper-based questionnaire and were asked to return it within 1 week. Responses were then entered using a purpose-built, web-based, data-entry system. The SCG-FFQ data were analysed using the UK food composition tables.

### 2.7. Power calculation and statistical analysis

We estimated our sample size using an priori approach employing G*Power (version 3.1). Our calculation indicates that a total sample size of 46 participants (23 per experimental group) is adequate considering a two-tailed significance (α=0.05) for a within-between interaction with a medium effect size (f = 0.25) between baseline and follow-up and ensuring a 90% power.

Analysis was conducted according to CONSORT guidelines for randomised pilot and feasibility trials ^57^, with all participants randomised being included (n=52). Descriptive statistics were used to compare baseline characteristics of trial participants by allocated group using IBM SPSS Statistics software (Version 29.0.1.0). The Shapiro-Wilk test was used to test the normality of the data at baseline, and the significance of baseline differences between the two-intervention groups was determined using either the Mann-Whitney U test or the unpaired t-test with Welch’s correction in GraphPad Prism (version 10). The analysis for primary outcome measures was performed using a General Linear Model (GLM) with repeated measures including within-subject (time) and between-subject (intervention) factors. Although main effects are reported here, the results focused on testing interaction effects (i.e., time*intervention). *Post hoc* analysis using the Benjamini-Hochberg procedure was performed for multiple comparisons of group means (False Discovery Rate correction (FDR) with 5% threshold). GLM approach was conducted using GraphPad Prism (version 10). The ROUT method (q = 1%) was used to identify outliers. Transformation (e.g. log10) was performed where needed to achieve normality. Partially recorded objective sleep measures and those with low signal quality were removed from further analysis. Significance of the difference in dietary consumption between the two-intervention groups was determined using either the Mann-Whitney U test or unpaired t-test with Welch’s correction in GraphPad Prism (version 10).

For the gut microbiome analysis, Linear Discriminant Analysis Effect Size (LEfSe) was used to explain the differences among the biological groups and detect features with significant differential abundance among classes. M2IA software was used to investigate the relationship between the primary outcomes (subjective/objective sleep) and the gut microbiome shifts. ^58^

## 3. Results

### 3.1. Participants characteristics

A total of 90 individuals were assessed for eligibility, with 52 participants recruited and randomised to control (n=26) or saffron (n=26) intervention groups. Four participants were lost at baseline, the reason being loss of contact (n = 4; 7.1%). There were no differences in age, years of education, cognitive status as assessed by ACE III, or subjective/objective sleep scores. Fifty-two participants completed the trial, of which 71% were female (Table 1) and mainly of white British ethnic origin. The trial CONSORT flow diagram is provided in Figure 1. A total of 33 compliance spreadsheets were completed in the intervention and placebo group, and no side effects were reported.

**Figure 1:**
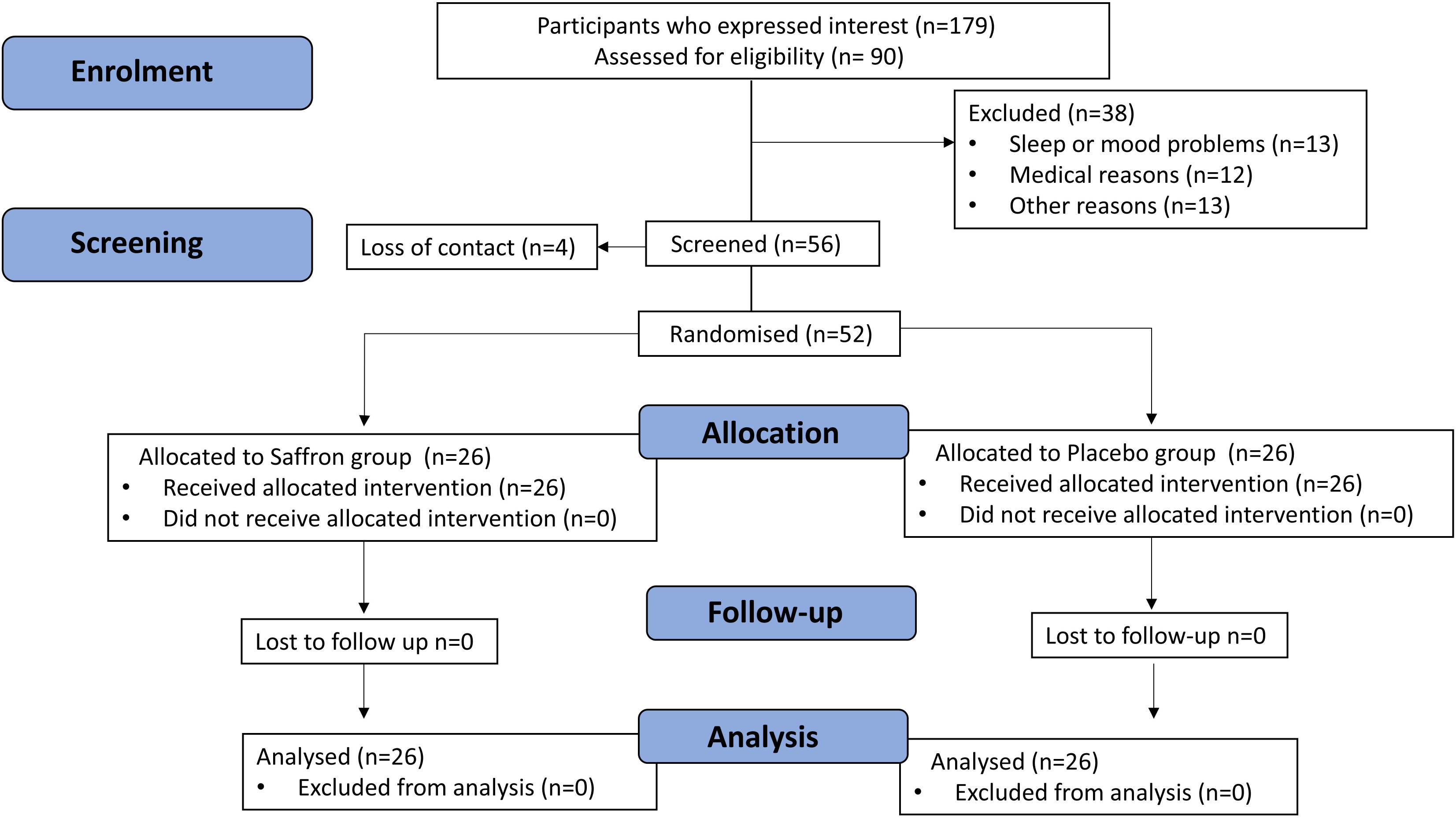
CONSORT flowchart diagram. A total of 52 participants were randomly assigned to either the saffron (n=26) or control group (n=26).

**Table 1:** Participant demographics.

| Characteristics | Placebo (n=26) | Saffron (n=26) | T-test |
| --- | --- | --- | --- |
|  | Mean (SD) | Mean (SD) | Sig. |
| Age (years) | 65.2 (7.3) | 66.3 (8.5) | 0.6 |
| Sex (M/F %) | 27/73 | 30/70 | 0.6 |
| Education (years) | 16.9 (4.4) | 18.7 (14.1) | 0.8 |
| BMI (KG/M<sup>2</sup>) | 26.1 (4.1) | 26.3 (4.6) | 0.8 |
| MINI ACE III | 28.5 (1.7) | 27.6.1 (2.7) | 0.2 |
| PHQ-9 (0-27) | 3.9 (4.2) | 4.5 (4.6) | 0.7 |
| GAD (0-21) | 2 (2.8) | 2.7 (3.3) | 0.4 |
| GHQ (0-28) | 12.5 (4.9) | 12.5 (5) | 0.98 |
| PANAS Positive (10-50) | 34.3 (10.9) | 30.2 (8.4) | 0.1 |
| PANAS Negative (10-50) | 17.2 (6.9) | 18.1 (7.9) | 0.6 |

Data are presented as mean (SD) or as stated (%). BMI: Body Mass Index; ACE III: Addenbrooke's Cognitive Examination-III; PHQ-9: Patient Health Questionnaire-9; GAD: General Anxiety Disorder; GHQ: General Health Questionnaire, PANAS: Positive and Negative Affect Schedule. Mann-Whitney test was used to test for baseline differences in non-normally distributed data; Unpaired t-test was used in normally distributed data (Shapiro-Wilk $p > 0.05$ ).

### 3.2. Primary outcomes: Sleep quality

#### 3.2.1. Effect of daily saffron supplementation on subjective sleep quality

A significant *time*\**intervention* interaction was observed in the PSQI global score with a reduction of 21.3% (F_1, 48_ = 5.82; p=0.02) in the saffron group over the 4-week intervention period when compared to an 8.6% increase from baseline in the placebo group (Figure 2A and Table 2). Furthermore, a significant interaction effect was observed in the sleep efficiency (%) (ratio calculated from the subjective hours slept and subjective hours in bed) PSQI sub score (F_1, 44_ = 4.291; p=0.04) which was increased by 5.4% in the saffron group and reduced by 14.7% in the placebo group at follow-up. A similar effect was seen in the PSQI sleep quality subcomponent score with a 36.36% increase in the placebo group and a 17.6% decrease in the saffron group (F_1, 48_ = 4.100; p=0.04). All these changes indicate a relative improvement in self-reported sleep in the saffron group compared to the placebo group during the intervention.

**Figure 2:**
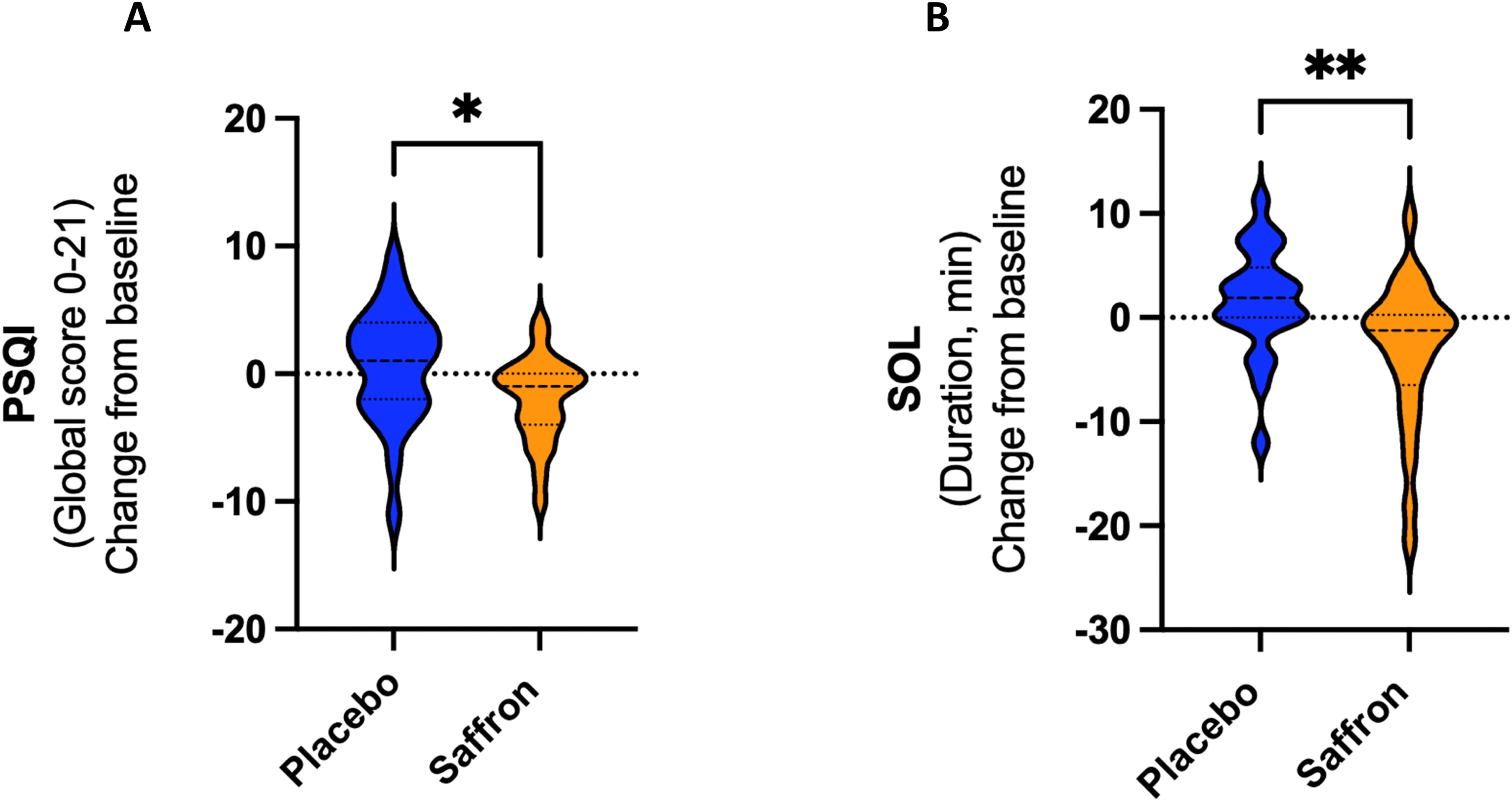
Violin plots representing differences in global PSQI score (A) and SOL duration (B) change from baseline, between the two intervention groups, with a greater reduction in the saffron group, indicating better improvement in sleep quality (p=0.017 and p=0.005, respectively). PSQI: Pittsburgh Sleep Quality Index; SOL: Sleep Onset Latency.

**Table 2:** Primary outcome results: Subjective and Objective sleep.

| Subjective sleep outcomes | Placebo BL<br>(n=26) | Saffron BL<br>(n=26) | Placebo FU<br>(n=26) | Saffron FU<br>(n=26) | Effects |  |  |
| --- | --- | --- | --- | --- | --- | --- | --- |
|  | Mean (SD) | Mean (SD) | Mean (SD) | Mean (SD) | p time (t) | p intervention (i) | t*i p interaction |
| PSQI global score | 8.1 (3.6) | 9.4 (2.6) | 8.8 (4.1) | 7.4 (3) | 0.2 | 0.9 | **0.02** |
| PSQI Sleep quality score | 1.1 (0.9) | 1.7 (0.7) | 1.5 (0.6) | 1.4 (0.7) | 0.9 | 0.1 | **0.05** |
| PSQI Sleep latency (in min) | 22.6 (16.8) | 23.5 (16.4) | 21.6 (12.3) | 13.3 (7.7) | **0.02** | 0.2 | 0.07 |
| PSQI Sleep duration (in min) | 361.9 (51.7) | 342 (54.1) | 383.8 (59.3) | 369.6 (72.3) | **0.03** | 0.2 | 0.7 |
| PSQI Sleep efficiency (in %) | 76.3 (13.4) | 69.9 (10.9) | 65.1 (23.7) | 73.7 (20.8) | 0.3 | 0.8 | **0.04** |
| PSQI Sleep disturbance score | 1.5 (0.6) | 1.6 (0.5) | 1.3 (0.5) | 1.6 (0.6) | 0.2 | 0.2 | 0.1 |
| PSQI Daytime dysfunction score | 1 (0.7) | 0.8 (0.8) | 0.7 (0.7) | 0.6 (0.6) | **0.02** | 0.4 | 0.8 |
| Epworth Sleepiness Scale (ESS) | 6.5 (5.1) | 8 (4.7) | 5.9 (4.2) | 6.5 (4.4) | **0.01** | 0.4 | 0.3 |
| Karolinska Sleepiness Scale (KSS) | 2.9 (1.7) | 2.1 (1.1) | 2.8 (1.5) | 2.7 (1.1) | 0.3 | 0.2 | 0.09 |
| Insomnia severity Index (ISI) | 8.7 (6.9) | 11.2 (6.7) | 11.1 (4.1) | 11.4 (5.6) | 0.3 | 0.3 | 0.3 |
| Objective sleep outcomes | Placebo BL<br>(n=21) | Saffron BL<br>(n=21) | Placebo FU<br>(n=21) | Saffron FU<br>(n=21) | p time (t) | p intervention (i) | t*i p interaction |
|  | Mean (SD) | Mean (SD) | Mean (SD) | Mean (SD) |  |  |  |
| Total sleep time (TST) (in min) | 418.6 (43.59) | 382.1 (62.8) | 418.6 (62) | 381.9 (45.6) | 0.9 | **0.01** | 0.6 |
| Sleep onset latency (SOL) (in min) | 14.7 (7.4) | 14.3 (6.9) | 17.5 (9.3) | 11.3 (5.2) | 0.9 | 0.06 | **0.03** |
| Wake after sleep onset (WASO) (in min) | 35.7 (14.7) | 26.1 (13.6) | 42.6 (17.5) | 24.5 (12.7) | 0.3 | **0.0004** | 0.2 |
| Wake duration (in min) | 66.7 (27.6) | 50.1 (25.4) | 70.8 (26.7) | 40.2 (20.9) | 0.9 | **0.005** | 0.008 |
| N1 sleep duration (in min) | 31.4 (9.1) | 27.8 (11.2) | 31.8 (9) | 27.2 (11.6) | 0.8 | 0.08 | 0.6 |
| **N1 %** | 7.9 (2.3) | 7.6 (2.4) | 7.6 (2.4) | 6.1 (1.6) | 0.08 | 0.1 | 0.06 |
| N2 sleep duration (in min) | 218.1 (34) | 191.9 (40.7) | 206.8 (41.7) | 179.5 (50.4) | 0.07 | **0.01** | 0.8 |
| **N2 %** | 51.9 (5.7) | 51.1 (6.8) | 50.1 (4.8) | 49.2 (6.4) | 0.06 | 0.7 | 0.8 |
| N3 sleep duration (in min) | 74.8 (20.2) | 70.8 (30.9) | 75.1 (17.5) | 73.1 (28.7) | 0.9 | 0.6 | 0.9 |
| **N3 %** | 18.4 (4.7) | 18.1 (6.6) | 19.1 (5.2) | 21.3 (7.7) | 0.053 | 0.6 | 0.1 |
| Rapid-eye-movement (REM) duration (in min) | 85.2 (25.9) | 79.4 (26.1) | 90.3 (31.2) | 80.9 (32.3) | 0.6 | 0.3 | 0.6 |
| **REM %** | 20.6 (5.3) | 21.1 (6.2) | 22.6 (5.9) | 19.4 (7.1) | 0.8 | 0.4 | **0.03** |
| Non-rapid-eye-movement (NREM) duration (in min) | 325.9 (31.5) | 299.6 (58.9) | 319.7 (45.6) | 290.4 (55.2) | 0.3 | **0.02** | 0.7 |
| Sleep efficiency (%) | 86.1 (5.9) | 88.1 (6.1) | 85.6 (5.3) | 90 (4.9) | 0.9 | 0.07 | 0.2 |
| Awakenings (in min) | 24.6 (5.5) | 21.3 (8.5) | 22.5 (3.6) | 20.5 (6.2) | 0.4 | 0.2 | 0.2 |
| Latency to persistent sleep (LtPS) (in min) | 17.5 (8.2) | 21 (12.4) | 23.7 (12.6) | 15.2 (7.1) | 0.8 | 0.3 | **0.003** |

Data are presented as mean (SD) or as *n* (%) for both baseline (BL) and follow-up (FU), adjusted for outliers (ROUT method, Q cutoff of 1%). P values are indicated for the two main effects (i.e., time and intervention) and the interactions, as derived from repeated measures GLM.

Several main effects of time were observed on daytime dysfunction (F_1, 48_ = 5.980; p=0.02), sleep duration (F _1, 46_ = 4.704; p=0.03) and sleep latency (F_1, 41_ = 5.434; p=0.02) PSQI sub-scores (Table 2). In addition, a main effect of time was observed on the ESS score (F_1, 48_ = 6.607; p=0.01) with a 9.2% reduction in the placebo group and an 18.7% reduction in the saffron group. No significant effects were observed in the Karolinska sleepiness scale (KSS) and the insomnia severity index (ISI).

#### 3.2.2. Effect of daily saffron supplementation on objective sleep quality

A total of 590 full night records were analysed. A significant *time*intervention* was observed for Latency to Persistent Sleep (LtPS) (F_1, 37_ = 9.777; p=0.003), with scores decreasing significantly in the saffron group (-27.6%) but increasing in the placebo group (+35.4%). Furthermore, sleep onset latency (SOL, min) decreased by 21% in the saffron group while it increased by 19.05% in the placebo group (F_1,38_ = 4.928, p=0.03) (Figure 2B). In addition, a significant *time*intervention* interaction was observed for REM sleep (%) increasing by 9.7% in the placebo group and a decreasing by 8.1% in the saffron group (F_1,40_ = 5.312; p=0.03) (Table 2).

Furthermore, a main effect of intervention was observed in the wakefulness after sleep onset (WASO, min) which was significantly increased in the placebo group (+19.33%) but reduced by 6.1% in the saffron group (F_1,40_ = 14.92; p=0.0004). A main effect of intervention in total sleep time (TST; min) (F_1,43_ = 6.744; p=0.01), in time spent in N2 (F_1,44_ = 6.486; p=0.01), and overall NREM (F_1,44_ = 5.872; p=0.02) and wake duration (min) (F_1,43_ = 8.665; p=0.0052) were also observed (Table 2).

### 3.3. Gut microbiome

#### 3.3.1. FFQ analysis in the microbiome sub-cohort

Table 3 presents the results from the FFQ-SCG at baseline. Significant differences were observed in the treatment groups with the placebo group consuming more energy (F_8,12_ = 1.336; p=0.03), more proteins (F_8,12_ = 1.153; p=0.01), more fibres (AOAC) (F_13,10_ = 1.248; p=0.03) and more total sugars (F_7,12_ = 1.856; p=0.02) than the saffron group. In addition, a significant difference was also observed for flavones (mg) intake which were higher in the placebo group (F_12, 9_ = 6.137; p=0.008). The average intake of total flavonoid and carotenoid content was similar between the two groups. However, alcohol intake was reported to be higher in the saffron than in placebo group (F_7,8_ = 18.08; p=0.02).

**Table 3:**
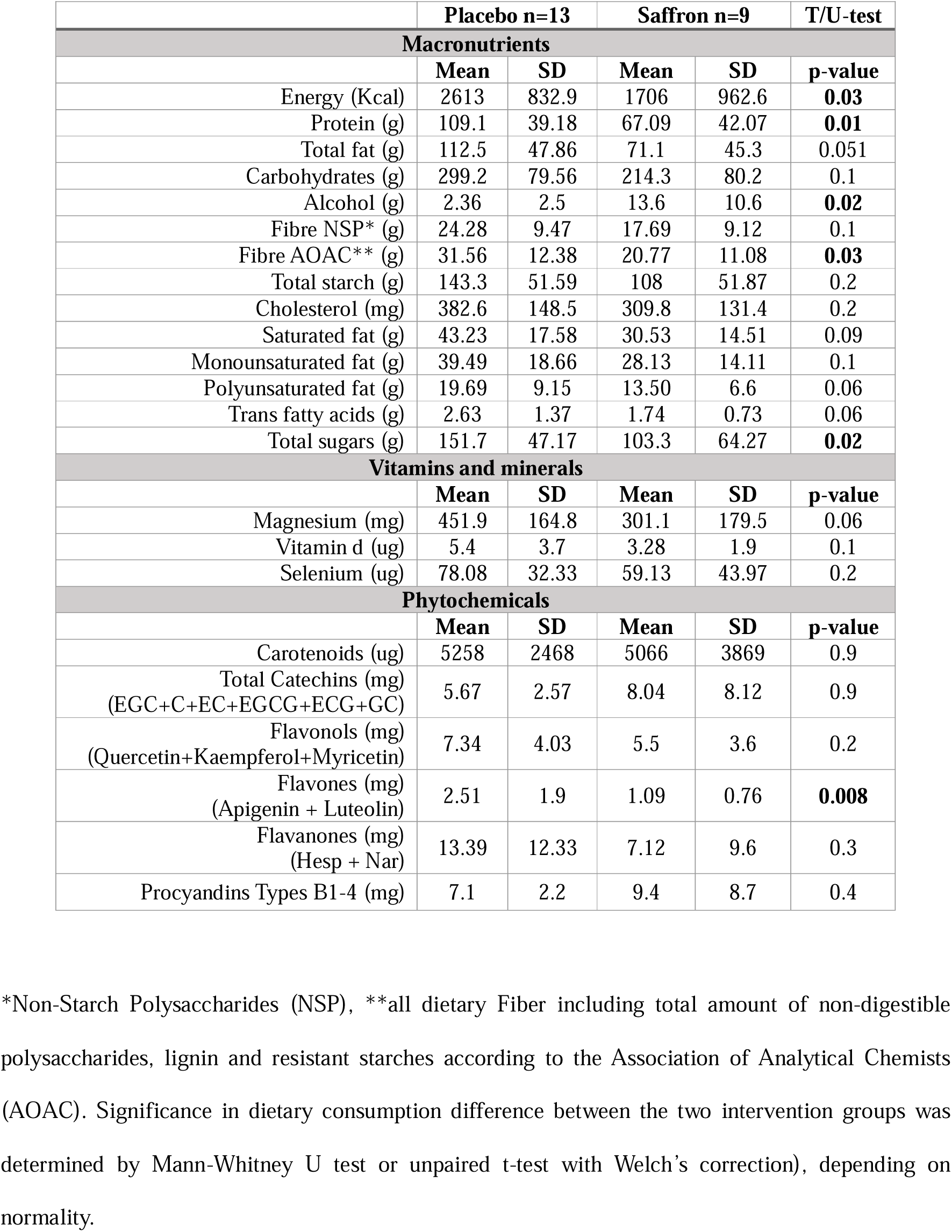
Group differences in macronutrient intake at baseline, as measured by the Food Frequency Questionnaire (FFQ) within the microbiome cohort(n=22).

#### 3.3.2. 4-week daily saffron supplementation affected the microbiome speciation

After demonstrating a beneficial effect of saffron on both subjective and objective sleep measures, we next sought to investigate whether the gut microbiota was modified by the saffron treatment. Compared to the placebo group at follow-up, saffron intake led to an enrichment of Bacteroidota (class Bacteroidia) and Verrucomicrobiota (class Verrucomicrobiae) and a reduction in Firmicutes (class Bacilli (46%) and Gammaproteobacteria (29%). Furthermore, saffron intake decreased the Firmicute/Bacteroidetes ratio when compared to placebo after 4 weeks of supplementation (Figure 3A and Supplementary Table S1-S3). Saffron intake did not affect alpha diversity as assessed by Chao1 (p = 0.8) and Shannon diversity indices (p=0.4) (Figure 3B) nor beta diversity as measured by Bray-Curtis distance (PERMANOVA p=0.964; Figure 3C).

**Figure 3:**
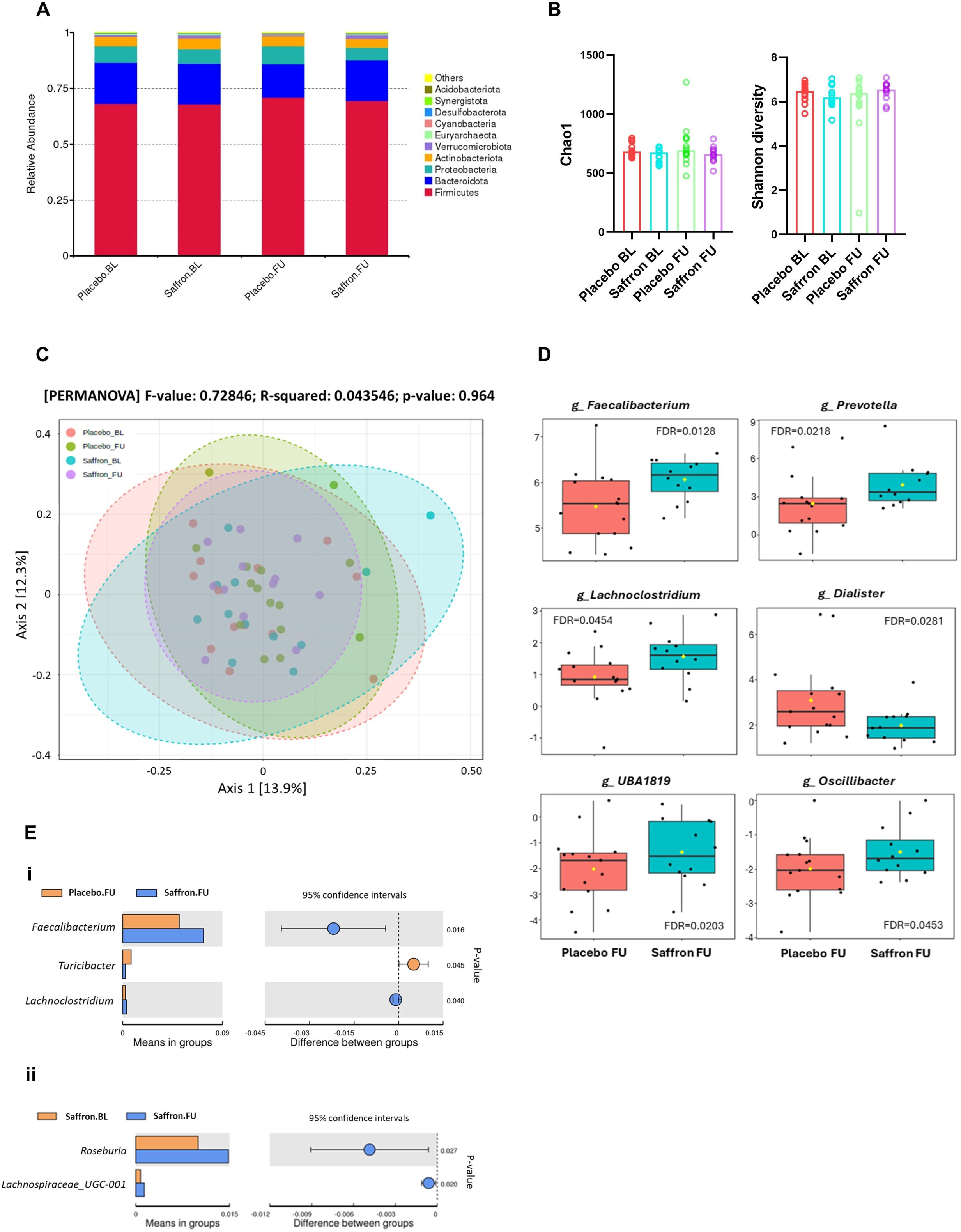
Impact of saffron on microbiota diversity and abundance. A) Relative abundance of microbiota at the phylum level. B) Alpha diversity as analysed using chao1 and Shannon diversity metrics. C) PCoA of beta diversity measured by Bray-Curtis analysis. D) Linear discriminant analysis of effect size analysis (LEfSe; LDA>2, FDR<0.05) comparing placebo and saffron groups at follow-up at the genus level. E) Univariate analysis using the Wilcoxon rank-sum test comparing (i) placebo and saffron at follow-up or (ii) the Wilcoxon signed-rank test for comparing the saffron group only at baseline and follow-up. BL: baseline; FU: follow-up.

We then examined whether the relative abundance of any taxa might differ between these groups, using a linear discriminant analysis of effect size analysis (LEfSe; LDA>2, FDR<0.05). At the genus level *Prevotella, Lachnoclostridium, UBA1819, Oscillibacter* and *Faecalibacterium* were increased, while *Dialister* decreased post-intervention in the saffron group (Supplementary Table S4 and Figure 3D). Using the Wilcoxon rank-sum test we observed a small increase in *Lachnosclotridium* (p=0.04) and a slight reduction in *Turicibacter* (p=0.04) (Figure 3Ei). Saffron also significantly increased abundance of *Roseburia* and *Lachnospiraceae_UGC-001* (p=0.02) (Figure 3Eii) at follow-up.

#### 3.3.3. Saffron induced changes in gut microbiome are correlated with changes in sleep quality

We questioned whether the changes in gut microbiome observed following saffron intake were correlated with changes in both subjective and objective sleep measures. At the genus levels, *Oscillibacter* was positively correlated with sleep efficiency (%) component of the PSQI (r = 0.63, p= 0.0007) and *UBA1819* negatively with sleep latency (min) of the PSQI (r = -0.49, p= 0.041) (Figure 4A).

**Figure 4:**
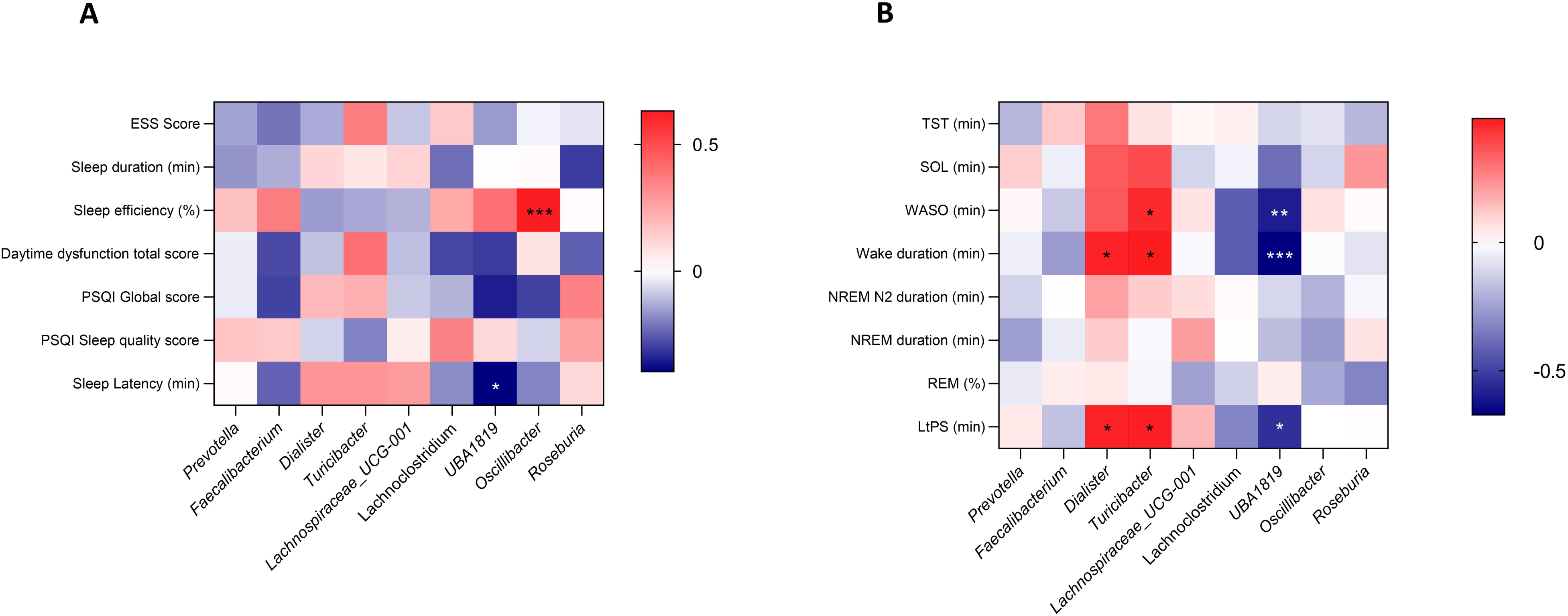
Heat map showing correlation results between (A) subjective and (B) objective sleep parameters and bacterial abundance at the genus level in the saffron versus placebo groups. P values for significance are presented as p<0.05 (*), p<0.01 (**) and p≤ 0.001 (***).

In the objective sleep dimension, latency to persistent sleep (LtPS, min) positively correlated with *Dialister* and *Turicibacter,* (r= 0.469 p=0.031 and r =0.474, p=0.029), but negatively with *UBA1819* (r = -0.545, p=0.01). *Dialister* and *Turicibacter* also positively correlated with wake duration (min), (r = 0.468, p=0.032 and r = 0.484, p=0.026) respectively. In addition, *Turicibacter* was also positively correlated with wake after sleep onset (WASO, min) (r= 0.449, p= 0.04). Negative correlations are also observed between *UBA1819* and wakefulness after sleep onset (WASO, min) (r=-0.595, p= 0.004), and wake duration (min) (r=-0.672, p=0.0008) (Figure 4B).

## Discussion

Building on emerging insights into the interaction between sleep, gut, and brain health, this study aimed at evaluating the effect of a patented standardised saffron extract (30mg per day for 4 weeks) not only on parameters of sleep quality (objective and subjective), but also on gut microbiome composition, in older adults with sleep complaints. Saffron supplementation over four weeks was associated with improvements in overall sleep quality. This is evidenced by a significant decrease in the PSQI global score, as well as improvement in the PSQI-derived sleep quality score and sleep efficiency % in saffron group compared to the control. The 21% reduction in PSQI score represents a substantial improvement in overall sleep quality, with the possibility of shifting from “poor sleep quality” to “better sleep quality”, and this improvement in sleep quality was furtherly validated by objective measures.

Objective sleep assessment using the Dreem device further indicated that the intervention had a significant impact on sleep quality, as evidenced by a reduction in sleep onset latency (SOL) and persistent sleep latency (LtPS), as well as in the time spent awake after sleep onset (WASO) and the duration of wakefulness. These changes reflect an overall improvement in sleep initiation and maintenance, which are critical aspects of sleep quality often adversely affected by insomnia symptoms and sleep disorders. The saffron group showed a slight decrease in the proportion of REM sleep at follow-up (19.4%) compared to baseline (21.1%), while the control group showed a slight increase in REM sleep at follow-up (22.6%) compared to baseline (20.6%). These different trajectories were supported by a marginally significant interaction between time and intervention. Although the cause of the reduction in REM sleep in the saffron group is unclear, the simultaneous increase in N3, associated with a slight decrease in N1 and no change in N2, suggests that the effect may be due to higher homeostatic sleep pressure in the saffron group at follow-up. This hypothesis is in line with other evidence we report showing a decrease in sleep latency after saffron treatment. In addition, saffron has been shown to have antidepressant effects, consistent with an increase in serotonergic neurotransmission, which may also contribute to the relative reduction in REM sleep.^59, 60^ However, we did not observe any changes in the ISI score in our cohort, reported to be positively modulated by saffron consumption by others ^61^, nor in the Karolinska Sleepiness Scale (KSS). These discrepancies may be explained by the fact that the KSS scale specifically measures daytime sleepiness over a short period (in the last 5-10 minutes) on a given day, whereas ESS measures sleepiness over a longer period and is less impact by short-term fluctuations. The lack of change in the ISI may be due to the fact that the baseline ISI levels in our cohort were at the lower end of the “insomnia subthreshold” (scores below 14). The subjective improvement in sleep quality observed in the questionnaires has already been reported by others ^61, 62^, but to our knowledge, we are the first to report positive changes in objective sleep measures (EEG).

The Dreem system allowed for an automatic characterisation of sleep architecture as validated against gold standard polysomnography ^51^, with the added advantage of monitoring these changes in the natural environment of the participants. We found positive effects of saffron intake on SOL, LtPS and WASO. High SOL and LtPS values reflect overall sleep issues, particularly sleep initiation difficulty, reminiscent of sleep onset insomnia. In contrast high WASO values indicate frequent or prolonged awakenings during the night and are a sign of discontinuous sleep with direct impact on the restorative function of sleep and reminiscent of sleep maintenance insomnia. These negative effects on sleep quality are often associated with impaired cognitive functions, mood disturbances and memory issues. ^61, 63^ Similarly, high LtPS values are indicative of poor sleep quality and are typically associated with reduced sleep efficiency, offering a complementary and more precise measure of SOL. ^64^

In general, people with insomnia complaints tend to underestimate the subjective duration of their sleep compared to that measured objectively by polysomnography. ^65^ Nevertheless, in our study, indications of the positive effects of saffron on sleep initiation and maintenance were corroborated by objectively measured sleep data. The convergence of the relatively large collection of objective sleep measurement data from Dreem recordings with the subjective PSQI results provides strong pilot evidence of the positive impact of this intervention on sleep quality.

Saffron consumption also led to significant changes in the gut microbiome, notably increasing beneficial bacteria such as in *Faecalibacterium*, *Prevotella*, *Roseburia* and *Lachnospiraceae_UGC-001* abundance. While gummy supplements may cause minor microbiome changes through prebiotic effects from pectin and maltitol [48], other compounds like sorbitol and stevia in small amounts are unlikely to have significant impacts. The placebo group received gummy supplements containing all the similar ingredients, ensuring a controlled comparison. Among the modulated bacteria, *Roseburia*, *Faecalibacterium* and *Lachnospiraceae* are known producers of butyrate. ^66–68^ Butyrate, a short-chain fatty acid generated by gut bacteria through the fermentation of dietary fibres, has been reported to act as sleep-promoting signal in rodent models upon oral or intraportal administration, indicating sleep-inducing sensory mechanism via gut microbes. ^69^ Therefore, increase in *Faecalibacterium*, *Lachnospiraceae* and *Roseburia* observed in the saffron group could potentially mediate the positive changes in sleep parameters via SCFAs production and gut-brain axis interactions, supporting the link between microbiome composition and overall sleep quality. In addition, the fact that dietary fibre intake was significantly higher in the placebo group further reinforces the hypothesis that saffron may be responsible for the positive changes observed in the gut microbiome. Similarly, the intake of phytochemicals (including carotenoids), vitamins or minerals (including magnesium (Mg) and vitamin D) that could have affected sleep parameters and interfered with the intervention treatment was no different in the participants’ usual diet. These positive changes in the profile of the intestinal microbiome demonstrate the prebiotic effects of saffron, hence playing a crucial role in intestinal homeostasis in addition to the known biological and neuroprotective effects of saffron’s apocarotenoids. ^70^

In our study, we reported that the genus *Oscillibacter* was strongly positively correlated with the sleep efficiency component of the PSQI in the subjective sleep measures. This genus of Firmicutes in the family *Oscillospiraceae* increased in the saffron group, has been previously associated with a decreased risk of insomnia and improvement in duration ^71^, which can be credited with improving sleep efficiency. Still in the subjective sleep measures, *UBA1819*, family Ruminococcaceae, which was also increased in saffron, was found to be negatively correlated with PSQI sleep latency (min), which was reduced in saffron. This family was reported to modulate sleep and to be inversely associated with chronic insomnia and linked to better sleep efficiency ^72^ (potentially also mediating glucose and lipid metabolism). ^73, 74^ In parallel, in the objective sleep measures, this genus was negatively correlated with measures of sleep fragmentation variables (LtPS, WASO and wake duration), that were decreased post-saffron supplementation. Reduction in sleep disruption variables are contributing factors to the improvement in sleep efficiency and quality, reinforcing the potential mediatory effect between Ruminococcaceae familly and sleep quality. Furthermore, in the objective sleep measures, those measures of sleep fragmentation were positively associated with *Turicibacter* (family Erysipelotrichaceae) and *Dialister* (family Veillonellaceae), genera that are both decreased in saffron, but in a very subtle manner for *Turicibacter*. Although there is limited available evidence regarding sleep and the abundance of those bacteria, *Dialister* genus was reported in both long and short sleepers but in African-origin adults ^75^, and *Turicibacter* genus was found to be decreased in young children with low total sleep time. ^76^

This study proposes for the first time potential associations between saffron supplementation, complementary measures of sleep quality and gut microbiome, highlighting several pathways between sleep and gut health, and validating the previous murine findings ^77–80^ along with confirming previous research exploring the effects of saffron on sleep improvement. ^28, 29, 61^

### Strengths and limitations

An important strength of our study was the use of the wearable sleep EEG (DREEM 3) device, which proved both feasible and acceptable to participants, with an average sleep recording of over 6 nights at both baseline and follow-up, and an average of 85% overall accuracy for sleep recordings (Dreem 3 headband), thus representing a useful technical approach to measuring objective sleep parameters in the home environment. While allowing for objective sleep assessment with higher ecological validity compared to in-lab polysomnography, the accuracy of Dreem 3 may not match that of PSG, particularly for specific sleep parameters. Indeed, the study effectively measures total sleep duration and efficiency, but other metrics in the objective sleep stages such as REM and NREM sleep might lack precision. ^81^

The innovative design of this study enabled a comprehensive assessment of the links between sleep, saffron and the gut microbiome, using complementary measures to compare and contrast sleep both subjectively and objectively. Although a pilot study, our sample size (52 participants) was sufficient to obtain statistical significance and to provide a valuable baseline for future research. Potential limitations include demographic biases such as sex (71 % female participants), and absence of mechanistic insights into saffron’s effects with no characterisation of saffron’s derived systemic metabolites.

The lack of change in the Insomnia Severity Index (ISS) and Karolinska Sleepiness Scale (KSS) score may suggest the need to consider the effects of distinct psychological and psychiatric comorbidities that can affect sleep perception and have implications for social and health outcomes, key factors in older populations. This underscores the complexity of sleep and its impact on health. Elderly people with sleep disorders represent a population in crucial need of sleep quality improvement, as they are more susceptible to the deleterious effects of sleep deficits on cognitive function and brain health, due to higher baseline cognitive vulnerability and reduced cognitive reserve.

## Conclusion

Our results indicate that short-term daily dietary intervention (4weeks) using a standardised saffron extract (30mg/day) can have positive impacts on both subjective and objective sleep quality in older adults with sleep complaints. This effect may be partly associated with changes in gut microbiota speciation, highlighting the potential of saffron in alleviating sleep disturbances in older adults by targeting the gut microbiome. While promising, these findings require further validation within a larger, diverse sample including a more balanced gender distribution.

## Author contributions

**Conceptualization**: ASL, DV. **Funding acquisition**: ASL, DV. **Investigation**: LL, AD, AS, TS. **Data curation**: LL, AD, VJ, ASL, DV. **Project administration**: ASL, DV, JT. **Resources**: LP, DG. **Formal analysis**: LL, AD, AS, TS, ASL, DV. **Writing – original draft**: LL, MGP, SMcA, ASL, DV. **Review and editing**: LL, AD, AS, VJ, SMcA, LP, DG, MGP, JT, TS, SS, MM, MH, ASL, DV.

## Data availability

The 16S rRNA gene sequence data have been deposited in the NCBI BioProject database (https://www.ncbi.nlm.nih.gov/bioproject/) under accession number PRJNA998744 (https://dataview.ncbi.nlm.nih.gov/object/PRJNA998701). Other original data will be made available from the corresponding author on reasonable request.

## Acknowledgements

This project was supported by the UK Research & Innovation (UKRI) Healthy Ageing Catalyst Awards (Grant ES/W006367/1). We would also like to thank the participants for their time and contribution to the study.

## References

1. W. H. Organisition, Journal, 2024.

2. Y. Hou, X. Dan, M. Babbar, Y. Wei, S. G. Hasselbalch, D. L. Croteau and V. A. Bohr, Ageing as a risk factor for neurodegenerative disease, Nat Rev Neurol, 2019, 15, 565–581.

3. A. M. V. Wennberg, M. N. Wu, P. B. Rosenberg and A. P. Spira, Sleep Disturbance, Cognitive Decline, and Dementia: A Review, Semin Neurol, 2017, 37, 395–406.

4. J. Li, M. V. Vitiello and N. S. Gooneratne, Sleep in Normal Aging, Sleep Med Clin, 2018, 13, 1–11.

5. B. Guarnieri and S. Sorbi, Sleep and Cognitive Decline: A Strong Bidirectional Relationship. It Is Time for Specific Recommendations on Routine Assessment and the Management of Sleep Disorders in Patients with Mild Cognitive Impairment and Dementia, Eur Neurol, 2015, 74, 43–48.

6. L. Ferini-Strambi, C. Liguori, B. P. Lucey, B. A. Mander, A. P. Spira, A. Videnovic, C. Baumann, O. Franco, M. Fernandes, O. Gnarra, P. Krack, M. Manconi, D. Noain, S. Saxena, U. Kallweit, W. Randerath, C. Trenkwalder, I. Rosenzweig, A. Iranzo, M. Bradicich and C. Bassetti, Correction to: Role of sleep in neurodegeneration: the consensus report of the 5th Think Tank World Sleep Forum, Neurol Sci, 2024, 45, 1813.

7. J. Kaur, L. M. Fahmy, E. Davoodi-Bojd, L. Zhang, G. Ding, J. Hu, Z. Zhang, M. Chopp and Q. Jiang, Waste Clearance in the Brain, Front Neuroanat, 2021, 15, 665803.

8. L. Xie, H. Kang, Q. Xu, M. J. Chen, Y. Liao, M. Thiyagarajan, J. O’Donnell, D. J. Christensen, C. Nicholson, J. J. Iliff, T. Takano, R. Deane and M. Nedergaard, Sleep drives metabolite clearance from the adult brain, Science, 2013, 342, 373–377.

9. E. S. Musiek and D. M. Holtzman, Mechanisms linking circadian clocks, sleep, and neurodegeneration, Science, 2016, 354, 1004–1008.

10. F. P. Cappuccio, L. D’Elia, P. Strazzullo and M. A. Miller, Sleep duration and all-cause mortality: a systematic review and meta-analysis of prospective studies, Sleep, 2010, 33, 585–592.

11. A. A. da Silva, R. G. de Mello, C. W. Schaan, F. D. Fuchs, S. Redline and S. C. Fuchs, Sleep duration and mortality in the elderly: a systematic review with meta-analysis, BMJ Open, 2016, 6, e008119.

12. G. G. Alvarez and N. T. Ayas, The impact of daily sleep duration on health: a review of the literature, Prog Cardiovasc Nurs, 2004, 19, 56–59.

13. E. E. Jaqua, M. Hanna, W. Labib, C. Moore and V. Matossian, Common Sleep Disorders Affecting Older Adults, Perm J, 2023, 27, 122–132.

14. E. Koffel, S. Ancoli-Israel, P. Zee and J. M. Dzierzewski, Sleep health and aging: Recommendations for promoting healthy sleep among older adults: A National Sleep Foundation report, Sleep Health, 2023, 9, 821–824.

15. K. V. Johnson, Gut microbiome composition and diversity are related to human personality traits, Hum Microb J, 2020, 15, None.

16. S. M. Lee, M. M. Milillo, B. Krause-Sorio, P. Siddarth, L. Kilpatrick, K. L. Narr, J. P. Jacobs and H. Lavretsky, Gut Microbiome Diversity and Abundance Correlate with Gray Matter Volume (GMV) in Older Adults with Depression, Int J Environ Res Public Health, 2022, 19.

17. A. Chakrabarti, L. Geurts, L. Hoyles, P. Iozzo, A. D. Kraneveld, G. La Fata, M. Miani, E. Patterson, B. Pot, C. Shortt and D. Vauzour, The microbiota–gut–brain axis: pathways to better brain health. Perspectives on what we know, what we need to investigate and how to put knowledge into practice, Cellular and Molecular Life Sciences, 2022, 79.

18. P. Strandwitz, Neurotransmitter modulation by the gut microbiota, Brain Res, 2018, 1693, 128–133.

19. B. B. Williams, A. H. Van Benschoten, P. Cimermancic, M. S. Donia, M. Zimmermann, M. Taketani, A. Ishihara, P. C. Kashyap, J. S. Fraser and M. A. Fischbach, Discovery and characterization of gut microbiota decarboxylases that can produce the neurotransmitter tryptamine, Cell Host Microbe, 2014, 16, 495–503.

20. B. Neroni, M. Evangelisti, G. Radocchia, G. Di Nardo, F. Pantanella, M. P. Villa and S. Schippa, Relationship between sleep disorders and gut dysbiosis: what affects what?, Sleep Med, 2021, 87, 1–7.

21. M. Yue, C. Jin, X. Jiang, X. Xue, N. Wu, Z. Li and L. Zhang, Causal Effects of Gut Microbiota on Sleep-Related Phenotypes: A Two-Sample Mendelian Randomization Study, Clocks & Sleep, 2023, 5, 566–580.

22. M. G. Gareau, E. Wine, D. M. Rodrigues, J. H. Cho, M. T. Whary, D. J. Philpott, G. Macqueen and P. M. Sherman, Bacterial infection causes stress-induced memory dysfunction in mice, Gut, 2011, 60, 307–317.

23. L. Desbonnet, G. Clarke, A. Traplin, O. O’Sullivan, F. Crispie, R. D. Moloney, P. D. Cotter, T. G. Dinan and J. F. Cryan, Gut microbiota depletion from early adolescence in mice: Implications for brain and behaviour, Brain Behav Immun, 2015, 48, 165–173.

24. A. M. Taylor, S. V. Thompson, C. G. Edwards, S. M. A. Musaad, N. A. Khan and H. D. Holscher, Associations among diet, the gastrointestinal microbiota, and negative emotional states in adults, Nutr Neurosci, 2020, 23, 983–992.

25. A. Fattorusso, L. Di Genova, G. B. Dell’Isola, E. Mencaroni and S. Esposito, Autism Spectrum Disorders and the Gut Microbiota, Nutrients, 2019, 11.

26. T. T. Huang, J. B. Lai, Y. L. Du, Y. Xu, L. M. Ruan and S. H. Hu, Current Understanding of Gut Microbiota in Mood Disorders: An Update of Human Studies, Front Genet, 2019, 10, 98.

27. H. Binks, E. V. G, C. Gupta, C. Irwin and S. Khalesi, Effects of Diet on Sleep: A Narrative Review, Nutrients, 2020, 12.

28. J. Lian, Y. Zhong, H. Li, S. Yang, J. Wang, X. Li, X. Zhou and G. Chen, Effects of saffron supplementation on improving sleep quality: a meta-analysis of randomized controlled trials, Sleep Med, 2022, 92, 24–33.

29. B. D. Pachikian, S. Copine, M. Suchareau and L. Deldicque, Effects of Saffron Extract on Sleep Quality: A Randomized Double-Blind Controlled Clinical Trial, Nutrients, 2021, 13.

30. R. Roustazade, M. Radahmadi and Y. Yazdani, Therapeutic effects of saffron extract on different memory types, anxiety, and hippocampal BDNF and TNF-alpha gene expressions in sub-chronically stressed rats, Nutr Neurosci, 2022, 25, 192–206.

31. F. Wauquier, L. Boutin-Wittrant, L. Pourtau, D. Gaudout, B. Moras, A. Vignault, C. Monchaux De Oliveira, J. Gabaston, C. Vaysse, K. Bertrand, H. Abrous, L. Capuron, N. Castanon, D. Vauzour, V. Roux, N. Macian, G. Pickering and Y. Wittrant, Circulating Human Serum Metabolites Derived from the Intake of a Saffron Extract (Safr’Inside(TM)) Protect Neurons from Oxidative Stress: Consideration for Depressive Disorders, Nutrients, 2022, 14.

32. M. Masaki, K. Aritake, H. Tanaka, Y. Shoyama, Z. L. Huang and Y. Urade, Crocin promotes non-rapid eye movement sleep in mice, Mol Nutr Food Res, 2012, 56, 304–308.

33. N. Umigai, R. Takeda and A. Mori, Effect of crocetin on quality of sleep: A randomized, double-blind, placebo-controlled, crossover study, Complementary Therapies in Medicine, 2018, 41, 47–51.

34. L. C. Beishon, A. P. Batterham, T. J. Quinn, C. P. Nelson, R. B. Panerai, T. Robinson and V. J. Haunton, Addenbrooke’s Cognitive Examination III (ACE-III) and mini-ACE for the detection of dementia and mild cognitive impairment, Cochrane Database Syst Rev, 2019, 12, CD013282.

35. K. Kroenke, R. L. Spitzer and J. B. Williams, The PHQ-9: validity of a brief depression severity measure, J Gen Intern Med, 2001, 16, 606–613.

36. R. L. Spitzer, K. Kroenke, J. B. Williams and B. Lowe, A brief measure for assessing generalized anxiety disorder: the GAD-7, Arch Intern Med, 2006, 166, 1092–1097.

37. D. P. Goldberg and V. F. Hillier, A scaled version of the General Health Questionnaire, Psychol Med, 1979, 9, 139–145.

38. J. R. Crawford and J. D. Henry, The positive and negative affect schedule (PANAS): construct validity, measurement properties and normative data in a large non-clinical sample, Br J Clin Psychol, 2004, 43, 245–265.

39. M. Jose Bagur, G. L. Alonso Salinas, A. M. Jimenez-Monreal, S. Chaouqi, S. Llorens, M. Martinez-Tome and G. L. Alonso, Saffron: An Old Medicinal Plant and a Potential Novel Functional Food, Molecules, 2017, 23.

40. D. Cerda-Bernad, L. Costa, A. T. Serra, M. R. Bronze, E. Valero-Cases, F. Perez-Llamas, M. E. Candela, M. B. Arnao, F. T. Barberan, R. G. Villalba, M. T. Garcia-Conesa and M. J. Frutos, Saffron against Neuro-Cognitive Disorders: An Overview of Its Main Bioactive Compounds, Their Metabolic Fate and Potential Mechanisms of Neurological Protection, Nutrients, 2022, 14.

41. T. Abu-Izneid, A. Rauf, A. A. Khalil, A. Olatunde, A. Khalid, F. A. Alhumaydhi, A. S. M. Aljohani, M. Sahab Uddin, M. Heydari, M. Khayrullin, M. A. Shariati, A. O. Aremu, A. Alafnan and K. R. R. Rengasamy, Nutritional and health beneficial properties of saffron (Crocus sativus L): a comprehensive review, Crit Rev Food Sci Nutr, 2022, 62, 2683–2706.

42. P. A. Jackson, J. Forster, J. Khan, C. Pouchieu, S. Dubreuil, D. Gaudout, B. Moras, L. Pourtau, F. Joffre, C. Vaysse, K. Bertrand, H. Abrous, D. Vauzour, J. Brossaud, J. B. Corcuff, L. Capuron and D. O. Kennedy, Effects of Saffron Extract Supplementation on Mood, Well-Being, and Response to a Psychosocial Stressor in Healthy Adults: A Randomized, Double-Blind, Parallel Group, Clinical Trial, Frontiers in Nutrition, 2021, 7.

43. C. Pouchieu, L. Pourtau, J. Brossaud, D. Gaudout, J.-B. Corcuff, L. Capuron, N. Castanon and P. Philip, Acute Effect of a Saffron Extract (Safr’InsideTM) and Its Main Volatile Compound on the Stress Response in Healthy Young Men: A Randomized, Double Blind, Placebo-Controlled, Crossover Study, Nutrients, 2023, 15.

44. D. J. Buysse, C. F. Reynolds, 3rd, T. H. Monk, S. R. Berman and D. J. Kupfer, The Pittsburgh Sleep Quality Index: a new instrument for psychiatric practice and research, Psychiatry Res, 1989, 28, 193–213.

45. C. H. Bastien, A. Vallieres and C. M. Morin, Validation of the Insomnia Severity Index as an outcome measure for insomnia research, Sleep Med, 2001, 2, 297–307.

46. M. W. Johns, A New Method for Measuring Daytime Sleepiness: The Epworth Sleepiness Scale, Sleep, 1991, 14, 540–545.

47. T. Akerstedt and M. Gillberg, Subjective and objective sleepiness in the active individual, Int J Neurosci, 1990, 52, 29–37.

48. E. Debellemaniere, S. Chambon, C. Pinaud, V. Thorey, D. Dehaene, D. Leger, M. Chennaoui, P. J. Arnal and M. N. Galtier, Performance of an Ambulatory Dry-EEG Device for Auditory Closed-Loop Stimulation of Sleep Slow Oscillations in the Home Environment, Front Hum Neurosci, 2018, 12, 88.

49. R. Cox, I. Korjoukov, M. de Boer and L. M. Talamini, Sound asleep: processing and retention of slow oscillation phase-targeted stimuli, PLoS One, 2014, 9, e101567.

50. R. K. Malhotra, D. B. Kirsch, D. A. Kristo, E. J. Olson, R. N. Aurora, K. A. Carden, R. D. Chervin, J. L. Martin, K. Ramar, C. L. Rosen, J. A. Rowley, I. M. Rosen and D. American Academy of Sleep Medicine Board of, Polysomnography for Obstructive Sleep Apnea Should Include Arousal-Based Scoring: An American Academy of Sleep Medicine Position Statement, J Clin Sleep Med, 2018, 14, 1245–1247.

51. P. J. Arnal, V. Thorey, E. Debellemaniere, M. E. Ballard, A. Bou Hernandez, A. Guillot, H. Jourde, M. Harris, M. Guillard, P. Van Beers, M. Chennaoui and F. Sauvet, The Dreem Headband compared to polysomnography for electroencephalographic signal acquisition and sleep staging, Sleep, 2020, 43.

52. C. Quast, E. Pruesse, P. Yilmaz, J. Gerken, T. Schweer, P. Yarza, J. Peplies and F. O. Glockner, The SILVA ribosomal RNA gene database project: improved data processing and web-based tools, Nucleic Acids Res, 2013, 41, D590–596.

53. S. Lee and D. K. Lee, What is the proper way to apply the multiple comparison test?, Korean J Anesthesiol, 2018, 71, 353–360.

54. J. L. Hollis, L. C. A. Craig, S. Whybrow, H. Clark, J. A. M. Kyle and G. McNeill, Assessing the relative validity of the Scottish Collaborative Group FFQ for measuring dietary intake in adults, Public Health Nutrition, 2016, 20, 449–455.

55. L. F. Masson, G. McNeill, J. O. Tomany, J. A. Simpson, H. S. Peace, L. Wei, D. A. Grubb and C. Bolton-Smith, Statistical approaches for assessing the relative validity of a food-frequency questionnaire: use of correlation coefficients and the kappa statistic, Public Health Nutrition, 2007, 6, 313–321.

56. J. L. Hollis, L. C. Craig, S. Whybrow, H. Clark, J. A. Kyle and G. McNeill, Assessing the relative validity of the Scottish Collaborative Group FFQ for measuring dietary intake in adults, Public Health Nutr, 2017, 20, 449–455.

57. S. M. Eldridge, C. L. Chan, M. J. Campbell, C. M. Bond, S. Hopewell, L. Thabane, G. A. Lancaster and P. c. group, CONSORT 2010 statement: extension to randomised pilot and feasibility trials, BMJ, 2016, 355, i5239.

58. Y. Ni, G. Yu, H. Chen, Y. Deng, P. M. Wells, C. J. Steves, F. Ju and J. Fu, M2IA: a web server for microbiome and metabolome integrative analysis, Bioinformatics, 2020, 36, 3493–3498.

59. J. Adrien, Neurobiological bases for the relation between sleep and depression, Sleep Medicine Reviews, 2002, 6, 341–351.

60. H. A. Hausenblas, D. Saha, P. J. Dubyak and S. D. Anton, Saffron (Crocus sativus L.) and major depressive disorder: a meta-analysis of randomized clinical trials, Journal of Integrative Medicine, 2013, 11, 377–383.

61. A. L. Lopresti, S. J. Smith, A. P. Metse and P. D. Drummond, Effects of saffron on sleep quality in healthy adults with self-reported poor sleep: a randomized, double-blind, placebo-controlled trial, J Clin Sleep Med, 2020, 16, 937–947.

62. S. K. Sadat Rafiei, S. Abolghasemi, M. Frashidi, S. Ebrahimi, F. Gharei, Z. Razmkhah, N. Tavousi, B. Mahmoudvand, M. Faani, N. Karimi, A. Abdi, M. Soleimanzadeh, M. Ahmadpour Youshanlui, S. F. Sadatmadani, R. Alikhani, Y. Pishkari and N. Deravi, Saffron and Sleep Quality: A Systematic Review of Randomized Controlled Trials, Nutr Metab Insights, 2023, 16, 11786388231160317.

63. A. L. Lopresti, S. J. Smith and P. D. Drummond, An investigation into an evening intake of a saffron extract (affron(R)) on sleep quality, cortisol, and melatonin concentrations in adults with poor sleep: a randomised, double-blind, placebo-controlled, multi-dose study, Sleep Med, 2021, 86, 7–18.

64. K. Kawai, K. Iwamoto, S. Miyata, I. Okada, M. Ando, H. Fujishiro, M. Ando, A. Noda and N. Ozaki, LPS and its relationship with subjective-objective discrepancies of sleep onset latency in patients with psychiatric disorders, Sci Rep, 2023, 13, 22637.

65. A. G. Harvey and N. K. Tang, (Mis)perception of sleep in insomnia: a puzzle and a resolution, Psychol Bull, 2012, 138, 77–101.

66. G. L. Hold, A. Schwiertz, R. I. Aminov, M. Blaut and H. J. Flint, Oligonucleotide probes that detect quantitatively significant groups of butyrate-producing bacteria in human feces, Appl Environ Microbiol, 2003, 69, 4320–4324.

67. S. H. Duncan, G. L. Hold, A. Barcenilla, C. S. Stewart and H. J. Flint, Roseburia intestinalis sp. nov., a novel saccharolytic, butyrate-producing bacterium from human faeces, Int J Syst Evol Microbiol, 2002, 52, 1615–1620.

68. M. Vital, A. C. Howe and J. M. Tiedje, Revealing the bacterial butyrate synthesis pathways by analyzing (meta)genomic data, mBio, 2014, 5, e00889.

69. E. Szentirmai, N. S. Millican, A. R. Massie and L. Kapas, Butyrate, a metabolite of intestinal bacteria, enhances sleep, Sci Rep, 2019, 9, 7035.

70. S. F. Omidkhoda and H. Hosseinzadeh, Saffron and its active ingredients against human disorders: A literature review on existing clinical evidence, Iran J Basic Med Sci, 2022, 25, 913–933.

71. Y. Li, Q. Deng and Z. Liu, The relationship between gut microbiota and insomnia: a bi-directional two-sample Mendelian randomization research, Front Cell Infect Microbiol, 2023, 13, 1296417.

72. R. P. Smith, C. Easson, S. M. Lyle, R. Kapoor, C. P. Donnelly, E. J. Davidson, E. Parikh, J. V. Lopez and J. L. Tartar, Gut microbiome diversity is associated with sleep physiology in humans, PLoS One, 2019, 14, e0222394.

73. A. Eetemadi and I. Tagkopoulos, Methane and fatty acid metabolism pathways are predictive of Low-FODMAP diet efficacy for patients with irritable bowel syndrome, Clin Nutr, 2021, 40, 4414–4421.

74. K. Lull, R. K. Arffman, A. Sola-Leyva, N. M. Molina, O. Aasmets, K. H. Herzig, J. Plaza-Diaz, S. Franks, L. Morin-Papunen, J. S. Tapanainen, A. Salumets, S. Altmae, T. T. Piltonen and E. Org, The Gut Microbiome in Polycystic Ovary Syndrome and Its Association with Metabolic Traits, J Clin Endocrinol Metab, 2021, 106, 858–871.

75. N. Fei, C. Choo-Kang, S. Reutrakul, S. J. Crowley, D. Rae, K. Bedu-Addo, J. Plange-Rhule, T. E. Forrester, E. V. Lambert, P. Bovet, W. Riesen, W. Korte, A. Luke, B. T. Layden, J. A. Gilbert and L. R. Dugas, Gut microbiota alterations in response to sleep length among African-origin adults, PLoS One, 2021, 16, e0255323.

76. Y. Wang, M. van de Wouw, L. Drogos, E. Vaghef-Mehrabani, R. A. Reimer, L. Tomfohr-Madsen and G. F. Giesbrecht, Sleep and the gut microbiota in preschool-aged children, Sleep, 2022, 45.

77. M. G. Pontifex, E. Connell, G. Le Gall, L. Pourtau, D. Gaudout, C. Angeloni, L. Zallocco, M. Ronci, L. Giusti, M. Muller and D. Vauzour, Saffron extract (Safr’Inside) improves anxiety related behaviour in a mouse model of low-grade inflammation through the modulation of the microbiota and gut derived metabolites, Food Funct, 2022, 13, 12219–12233.

78. G. Singh, H. Brim, Y. Haileselassie, S. Varma, A. Habtezion, M. Rashid, S. R. Sinha and H. Ashktorab, Microbiomic and Metabolomic Analyses Unveil the Protective Effect of Saffron in a Mouse Colitis Model, Curr Issues Mol Biol, 2023, 45, 5558–5574.

79. G. Singh, Y. Haileselassie, A. R. Ji, H. T. Maecker, S. R. Sinha, H. Brim, A. Habtezion and H. Ashktorab, Protective Effect of Saffron in Mouse Colitis Models Through Immune Modulation, Dig Dis Sci, 2022, 67, 2922–2935.

80. S. Banskota, H. Brim, Y. H. Kwon, G. Singh, S. R. Sinha, H. Wang, W. I. Khan and H. Ashktorab, Saffron Pre-Treatment Promotes Reduction in Tissue Inflammatory Profiles and Alters Microbiome Composition in Experimental Colitis Mice, Molecules, 2021, 26.

81. Z. Liang and M. A. Chapa Martell, Validity of Consumer Activity Wristbands and Wearable EEG for Measuring Overall Sleep Parameters and Sleep Structure in Free-Living Conditions, J Healthc Inform Res, 2018, 2, 152–178.

